# Concentration-effect relationships of plasma caffeine on EEG delta power and cardiac autonomic activity during human sleep

**DOI:** 10.1101/2023.10.14.23297036

**Authors:** Diego M. Baur, Dario A. Dornbierer, Hans-Peter Landolt

## Abstract

Acute caffeine intake affects brain and cardiovascular physiology, yet the concentration-effect relationships on the electroencephalogram (EEG) and cardiac autonomic activity during sleep are poorly understood. To tackle this question, we simultaneously quantified the plasma caffeine concentration with ultra-high-performance liquid chromatography, as well as the EEG, heart rate and high-frequency (0.15-0.4 Hz) spectral power in heart rate variability (HR-HRV), representing parasympathetic activity, with standard polysomnography during undisturbed human sleep. Twenty-one healthy young men ingested in randomized, double-blind, cross-over fashion, 160 mg caffeine or placebo in a delayed, pulsatile-release caffeine formula at their habitual bedtime, and initiated a four-hour sleep opportunity 4.5 hours later. The mean caffeine levels during sleep exhibited high individual variability between 0.2 and 18.4 µmol/l. Across the first two NREM-REM sleep cycles, EEG delta (0.75-2.5 Hz) activity and heart rate were reliably modulated by waking and sleep states. Caffeine dose-dependently reduced delta activity and heart rate, and increased HR-HRV in NREM sleep when compared to placebo. The average reduction in heart rate equaled 3.24 ± 0.77 beats per minute. Non-linear statistical models suggest that caffeine levels above ∼ 7.4 µmol/l decreased EEG delta activity, whereas concentrations above ∼ 4.3 µmol/l and ∼ 4.9 µmol/l, respectively, reduced heart rate and increased HR-HRV. The findings provide quantitative concentration-effect relationships of caffeine, EEG delta power and cardiac autonomic activity and suggest increased parasympathetic activity during sleep after intake of caffeine.

## Introduction

An oral dose of caffeine is rapidly absorbed, easily crosses the blood brain barrier, and affects sleep when taken shortly before bedtime (Bonati et al. 1982; Gardiner et al. 2023). It is widely accepted that the effects of moderate caffeine intake on sleep are mediated by competitive antagonism of adenosine A_1_ and A_2A_ receptors in the central nervous system (Lazarus et al. 2019; Reichert et al. 2022). Apart from the typical changes in sleep architecture (Clark and Landolt 2017), acute caffeine consistently attenuates EEG activity in the delta range (< 4.5 Hz) in non-rapid-eye-movement (NREM) sleep (Carrier et al. 2009; Drake et al. 2013; Landolt et al. 1995; Van Dongen et al. 2001). Interestingly, several studies demonstrated that caffeine concentrations deemed too low to exert an acute pharmacological response are followed by reduced low-frequency activity (< 2 Hz) during subsequent sleep (Landolt et al. 1995; Landolt et al. 2004; Rétey et al. 2007). Thus, it is currently unknown whether caffeine concentrations below the limit of detection, an attenuated build-up of sleep pressure by blockading adenosine receptors during wakefulness, or active metabolites such as paraxanthine underpin caffeine’s effect on sleep intensity (Reichert et al. 2022).

Apart from regulating the adaptive homeostatic response to the prior duration of sleep and wakefulness on the sleep EEG, adenosine also contributes to the regulation of other major physiological processes. For example, by acting on all known subtypes of adenosine receptors, adenosine elicits a complex hemodynamic response that reflects separate effects on the myocardium, the vascular tone, and the sympathetic nervous system (for review see Riksen et al., 2011). Exogenous *adenosine* administration to conscious humans elicits a consistent increase in heart rate and systolic blood pressure, and a drop in diastolic blood pressure (Riksen et al. 2011). On the other hand, the effects of *caffeine* on heart rate and blood pressure are typically small and differ widely among studies (Turnbull et al. 2017). Acute exposure to caffeine doses contained in 2-3 cups of coffee (> 200-300 mg) typically stimulates a transient increase in systolic and diastolic blood pressure, whereas both bradycardia and tachycardia were reported (Crooks et al. 2019; Riksen et al. 2011; Temple et al. 2017; Turnbull et al. 2017). Methylxanthine intoxication induces hypotension and tachyarrhythmias (Whitsett, et al. 1984). The divergent findings are likely related to the different doses administered, but also reflect genetic predisposition, caffeine intake habits and health status of study participants, as well as other possible influences (Crooks et al. 2019; Green et al. 1996; Koenig et al. 2013). They may also suggest that the blockade of adenosine receptors only plays a minor role in mediating the actions of caffeine on the cardiovascular system (Fredholm et al., 2017). At least at higher doses, increased intracellular calcium, release of norepinephrine, and dopamine receptor sensitization constitute other likely mechanisms of action of caffeine on cardiovascular functions (Temple et al. 2017).

We recently reported the development of a pulsatile-release caffeine formulation to attenuate symptoms of sleep inertia after sleep restriction (Dornbierer et al. 2021). Following low-dose (160 mg) caffeine intake in an engineered capsule at habitual bedtime, we observed a mean caffeine plasma concentration of ∼ 5 µmol/l at the beginning of a 4-hour sleep opportunity. We used this unique data set, to simultaneously quantify the evolution of caffeine levels, sleep architecture and the sleep EEG, as well as heart rate and the high-frequency component of the heart-rate variability (HF-HRV) spectrum as a marker of parasympathetic activity during sleep. Using generalized additive models, we aimed at establishing the concentration-effect relationships between caffeine in blood plasma, EEG low-frequency activity and cardiac autonomic activity in NREM sleep.

## Methods

The study protocol was approved by the Cantonal Ethics Committee of the Canton of Zurich (BASEC: 2018-00533) and registered on ClinicalTrials.gov (Identifier: NCT04975360). All participants provided written informed consent according to the declaration of Helsinki.

### Participants

Twenty-two healthy young men (mean age: 23.82 ± 2.96 years) completed the study. They met the following inclusion criteria: habitual caffeine consumption of less than 4 regular units per day (coffee, tea, chocolate, cola, energy drinks); male sex to avoid the impact of menstrual cycle on physiology during sleep; age between 18–34 years; body-mass-index between 20-25; Epworth Sleepiness Score (ESS) below 10; habitual sleep onset latency below 20 min (self-rated); regular sleep–wake rhythm with bedtime between 10 pm and 1 am; absence of any somatic or psychiatric disorders; no acute or chronic medication intake; non-smoker; and no history of drug abuse (lifetime use < 5, with exception of occasional cannabis use);. The participants were instructed to abstain from illicit drugs and caffeine during the entire study, starting two weeks prior to the first experimental night until the end of the study (the day after the second experimental night). No alcohol was allowed 24 h before the experimental nights. The minimal wash-out period of caffeine between experimental nights was 7 days. Starting two weeks prior to the first experimental night and lasting until the end of the study, participants were instructed to keep a regular sleep-wake rhythm, consistent with the volunteers’ habitual bedtime. To verify adherence to the regular sleep and wake times, participants wore a rest-activity monitor on the non-dominant arm and kept a sleep–wake diary. All included participants showed roughly a 22:00-06:00 (n = 2), 23:00-07:00 (n = 17) or 00:00-08:00 (n = 2) rest period. To facilitate the readability of the manuscript, clock times in text and Figures refer to the majority of participants who adhered to the 23:00-07:00 sleep schedule.

### Study design

All participants spent two nights in the sleep laboratory, separated by at least one week. Upon arrival between 20:00-21:00, participants were informed in detail about the study protocol. Afterwards, the electrodes for polysomnographic recordings were carefully placed according to standard criteria (Berry et al. 2017) and the venous catheter for continuous blood collection was applied. To ensure sleep restriction and prevent unintended sleep before the scheduled sleep opportunity, the participants were constantly supervised and engaged in selected table games. At their habitual bedtime, i.e., 4.5 hours before a 4-hour sleep opportunity scheduled according to the sleep-wake habits of each participant during the two weeks prior to the study, we administered a time-controlled, pulsatile-release formula of caffeine or placebo (matched in appearance). The study followed a randomized, double-blind, placebo-controlled, cross-over design.

The pulsatile-release caffeine formula was previously described in detail (Dornbierer et al. 2021). In brief, 160 mg caffeine per capsule was dispersed in coating media and sprayed onto inert microcrystalline cellulose spheres, to obtain various layers consisting of caffeine and release-controlling polymers. The micropellets were then encapsulated into hydroxypropyl-methylcellulose capsules.

### Caffeine quantification

To monitor the caffeine pharmacokinetics during polysomnographically recorded sleep in the soundproof and climatized bedrooms of the human sleep research unit, we collected blood samples from the left antecubital vein at baseline (0.5 hours before capsule intake) and 1.5, 2.5, 3.5, 4.5, 5.5, 6.5, 7.5, 8.5, 9.5, 10.5, 13.5 and 17.5 hours after capsule administration. The venous catheter was connected to a blood-collection setup in an adjacent room (Heidelberger plastic tube extensions through the wall) (Dornbierer et al. 2021). Blood samples (4 ml, BD Vacutainer EDTA) were collected without disturbing the sleeping study participants. The intravenous line was kept patent with a slow drip (10 ml/h) of heparinized saline (1000 IU heparin in 0.9 g NaCl/dl; HEPARIN Bichsel; Bichsel AG, 3800 Unterseen, Switzerland). Blood samples were immediately centrifuged for 10 min at 2000 relative centrifugal force (RCF) and plasma samples were immediately put on ice until final storage at −80°C.

For the quantification of caffeine in plasma, we used an ultra-high performance liquid chromatography system coupled to a linear ion trap quadrupole mass spectrometer operated in positive electrospray ionization with scheduled multiple reaction monitoring. We previously reported all experimental and analytical procedures in detail (Dornbierer et al. 2021). Because of unreliable measurements, we needed to exclude the caffeine concentration data of one participant from the analyses (Dornbierer et al. 2021).

To estimate the caffeine concentration at timepoints of interest during sleep, we fitted the hourly data collected after capsule intake using polynomial regression (R^2^ = 0.936; R package stats v. 4.2.1). Subsequently the regression curve was used to interpolate caffeine concentration during all sleep stages and epochs.

### Polysomnography

We recorded sleep with dedicated polysomnographic amplifiers (Artisan^®^, Micromed, Mogliano, Veneto, Italy). The recording setup consisted of 10 EEG electrodes according to the 10–20 system (Fp1, Fp2, F3, F4, C3, C4, P3, P4, O1, 02), a bipolar electrooculogram (EOG), a submental electromyogram (EMG), and a 2-lead electrocardiogram (ECG). We marked the individual EEG electrode coordinates by cutting a few hairs at the electrode positions, to ensure that the electrodes were placed at the same place in both experimental conditions. We sampled the analogue signals with a frequency of 256 Hz, conditioned them by a high-pass filter (EEG: -3 dB at 0.15 Hz; EMG: 10 Hz; ECG: 1 Hz) and an antialiasing low-pass filter (-3 dB at 67.2 Hz), and stored the digitized data with a resolution of 256 Hz. Because of insufficient quality, we needed to exclude the EEG data of one participant from the analyses.

We analyzed the polysomnographic data in Rembrandt^®^ Datalab (Version 8; Embla Systems, Planegg, Germany). We visually scored waking and sleep stages in 30-s epochs according to the criteria of the American Academy of Sleep Medicine (Berry et al., 2017). We visually identified and excluded movement- and arousal-related artifacts from the analyses.

We identified NREM-REM sleep cycles according to the criteria proposed by Feinberg & Floyd (Feinberg and Floyd 1979) with the recently published R-package SleepCycles (v.1.1.4) (Blume and Cajochen 2021). We visually inspected the results and corrected them if necessary. All participants completed at least two NREM-REM sleep cycles during the 4-hour sleep opportunity.

We computed the EEG power spectra between 0-30 Hz derived from the C3-A2 derivation by a Fast-Fourier transform based on 4-s epochs (Hanning window, linear detrending, 50% overlap), resulting in a frequency resolution of 0.25 Hz. We averaged spectral power in wakefulness (W), NREM sleep (stages N1, N2 and N3) and REM sleep. To investigate the evolution of EEG delta activity across the first two NREM-REM sleep cycles, we subdivided individual NREM sleep episodes into 20 equal parts and individual REM sleep episodes into four equal parts, and then averaged across all individuals (Landolt et al. 1995).

### Heart rate analyses

We identified the R-peaks in the ECG signal using the R-package rsleep (v. 1.0.4) and the scipy python package (v.1.10.0), manually confirmed the results and corrected them if necessary. Using the R-package RHRV (v.4.2.6), we computed HRV measures in the frequency domain in all sleep states during the first two NREM-REM sleep cycles. With the Fast Fourier Transform (FFT) algorithm implemented in the RHRV package, we quantified the power spectral density of HRV in the following frequency bands: high frequency (HF, 0.15-0.4 Hz), low frequency (LF, 0.05-0.15 Hz), very low frequency (VLF, 0.03-0.05 Hz), ultralow frequency (ULF, < 0.03 Hz). Because of the uncertain interpretation of the low-frequency components in the HRV spectrum (Berntson et al. 1997), we restricted the analyses to HF-HRV.

### Statistical analyses

We based all analyses on the complete data set of 20 participants using R version 4.2.1 (R Core Team, 2018) and RStudio Version 2022.07.1-554 (RStudio, Inc.). We analyzed the data with linear mixed effects models (R-package lme4 v.1.1-30 and lmerTest v. 3.1-3) or two-sided, paired t-tests where specified (R package rstatix v.0.7.0). We included the fixed effects *‘cycle’* (1, 2), *‘state’* (W, N1, N2, N3, REM) and/or *‘condition’* (placebo, caffeine), as well as their interactions in the statistical models. When multiple measurements per subject were available, we added *‘study participant’* as random effect. We checked distribution of residuals, goodness of fit and assumptions in all tests and models. In all Figures, we present group means and 95% confidence intervals, based on 1000 bootstrap replicates (R-package boot v.1.3-28) (Efron and Tibshirani 1993). To compare the placebo and caffeine conditions when the ANOVA *‘condition’* term yielded a significant result, we computed general linear hypothesis tests (R-package multcomp v. 1.4-19), corrected for multiple comparison with the Benjamini-Hochberg procedure. We also computed Cohen’s d measures, to quantify the effect size of the statistical differences (R-package rstatix v. 0.7.0) (Cohen 1988).

We used generalized additive models, to analyze the change in EEG delta activity, heart rate and HF-HRV as non-linear function of the caffeine concentration (R-package mgcv v.1.8-40, splines v.4.2.1). We computed the range of significance by comparing the temporal evolution of the fitted smooth function in the caffeine and placebo conditions (R-package mgViz v.0.1.9, gratia v.0.7.3). All graphics were generated using R-packages ggplot2 (v.3.3.6) or ggpubr (v.0.4.0).

## Results

### Caffeine plasma concentration

The delayed, pulsatile-release of caffeine started shortly before the sleep opportunity, and the caffeine in plasma exhibited a mean maximal concentration of 9.60 ± 0.89 μmol/l (± SEM, n=19) 9.5 hours after capsule intake (Fig. 1A). At lights-off, i.e., at the beginning of the sleep opportunity, the mean caffeine concentration equaled 4.19 ± 0.82 μmol/l, with individual values varying between 0.14-10.20 μmol/l. At lights-on of scheduled sleep, the mean caffeine concentration equaled 8.00 ± 0.86 μmol/l, varying between 0.65-16.4 μmol/l (Fig. 1B). The increase in caffeine across the 4-hour sleep opportunity varied between 0.51-16.9 μmol/l per individual.

**Figure 1.**
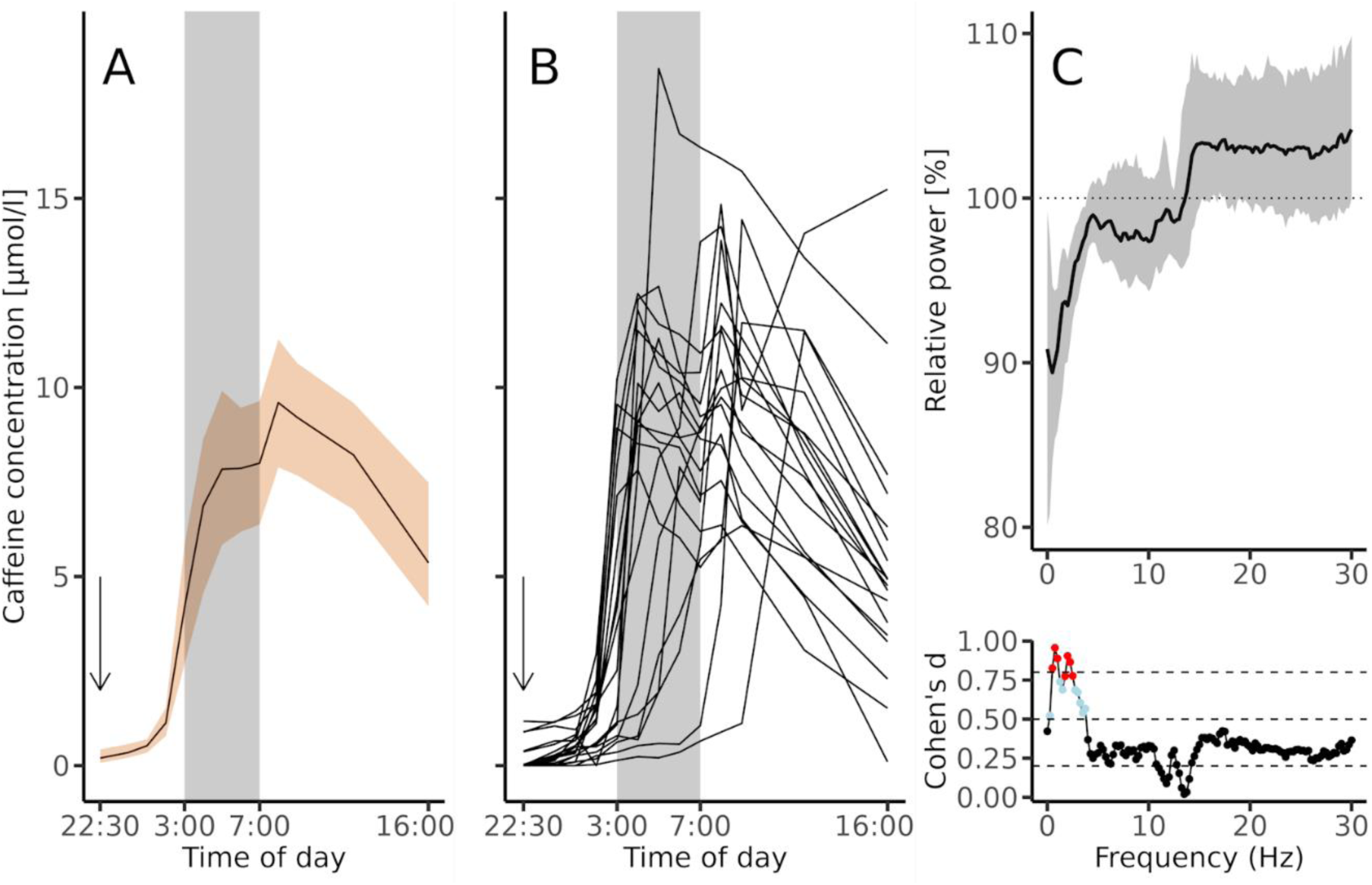
(**A**) Temporal evolution of the mean caffeine concentration in plasma (mean ± 95% confidence interval; n = 19). The x-axis refers to clock time. The vertical arrow indicates the time when the time-controlled, pulsatile-release formula containing 160 mg caffeine was ingested. Grey shading indicates the time-in-bed for sleep (**B**) Temporal evolution of individual caffeine kinetics of the 19 participants for whom caffeine data were available. (**C**) Relative EEG power spectra in NREM sleep (stages N1-N3). For each 0.25-Hz frequency bin between 0-30 Hz, mean power (± 95% confidence interval; n = 21) in the caffeine condition was expressed as a percentage of the corresponding value in the placebo condition (horizontal dashed lines at 100%). Bottom panel: Effect size of the difference between caffeine and placebo expressed as Cohen’s d. Red dots: p_FDR_ < 0.05; grey dots: p_uncorrected_ < 0.05; black dots: p_uncorrected_ > 0.05.

### Effects of caffeine on the EEG during sleep

The visually scored sleep variables in the first two NREM-REM sleep cycles in the placebo and caffeine conditions are summarized in supplementary Table S1. In both conditions, the participants quickly fell asleep and exhibited a high proportion of slow wave sleep. Except for the concentration-dependent, reduced time spent in N3 sleep after caffeine compared to placebo (69.7 ± 19.6 *vs.* 55.8 ± 19.5, p_FDR_ < 0.03) (also see supplementary Fig. S1), sleep architecture in both experimental conditions was comparable (p_all_ > 0.1).

Corroborating less visually scored N3 sleep, caffeine reduced EEG delta activity in NREM sleep. While power in all bins between 0.5-3.75 Hz was reduced, correction for multiple comparisons revealed that the reduction in the 0.75-2.5 Hz range was statistically significant (Fig. 1C). The caffeine intake reduced EEG activity in this frequency band with a large effect size when compared to placebo (average Cohen’s d 0.86 ± 0.02 [SEM]).

The evolution of EEG power in the 0.75-2.5 Hz band in the placebo and caffeine conditions across the first two NREM-REM sleep cycles is illustrated in Fig. 2A. Delta activity varied across wakefulness and sleep states (*‘state’*: F_4,180_ = 164.18, p < 0.0001). It decreased from the 1^st^ to the 2^nd^ NREM sleep episode (*‘cycle’*: F_1,60_ = 20.32, p < 0.0001) and was reduced after caffeine when compared to placebo (*‘condition’*: F_1,60_ = 7.52, p < 0.01). The mean effect size of the difference between the conditions equaled 0.28 ± 0.03 (SEM), indicating a small to medium effect. A decrease in 0.75-2.5 Hz activity was also present from the 1^st^ to the 2^nd^ REM sleep episode (*‘cycle’*: F_1,37.2_ = 6.66, p < 0.02), whereas the caffeine and placebo conditions did not differ (*‘condition’*: F_1,36.2_ = 3.83, p > 0.058).

**Figure 2.**
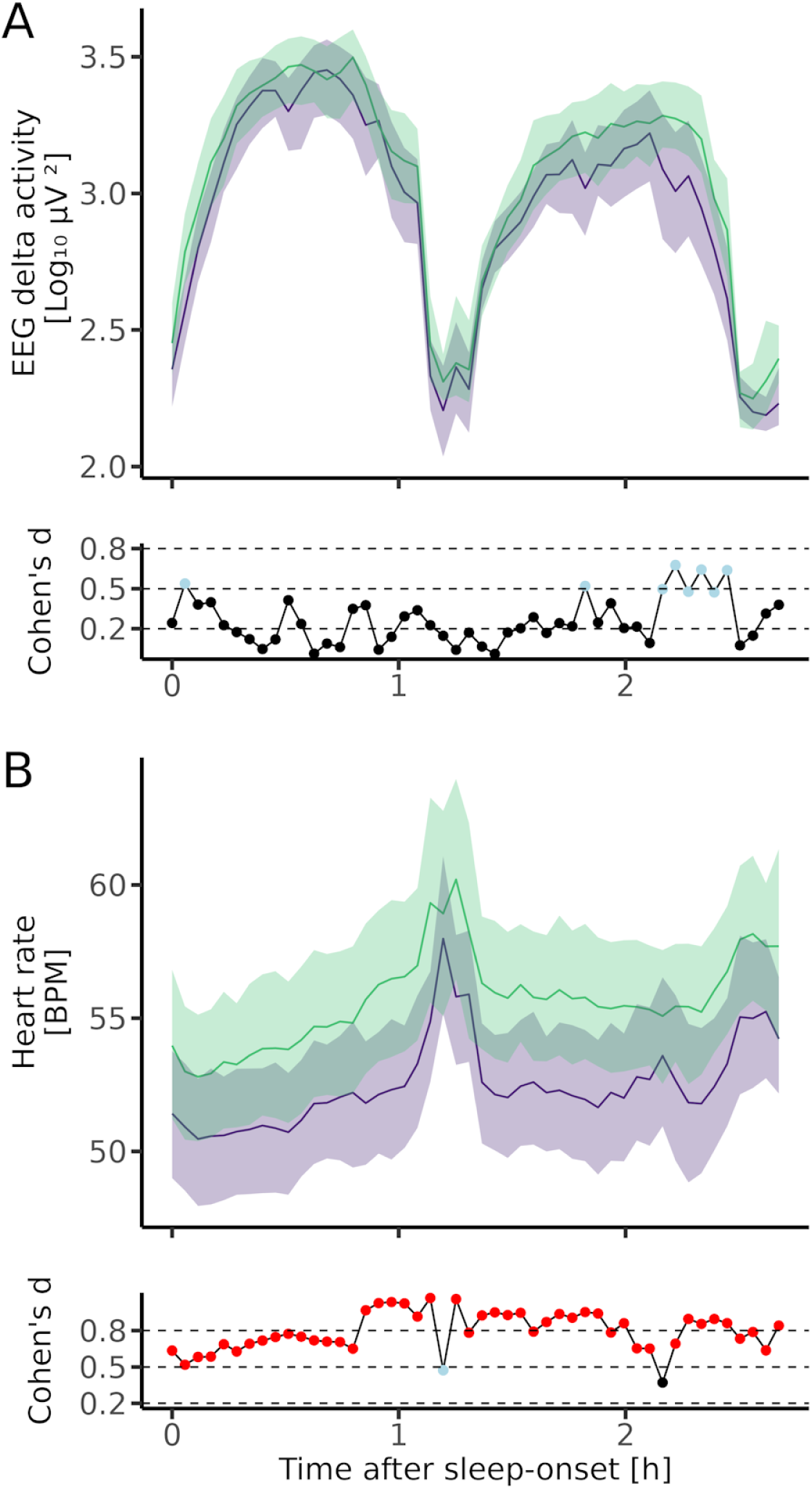
(**A**) Time course of EEG delta activity (0.75-2.5 Hz) across the first two NREM-REM sleep episodes in the placebo (green) and caffeine (purple) conditions (mean ± 95% confidence intervals; n = 21). Individual NREM sleep episodes were subdivided into 20 and individual REM sleep episodes into 4 equal time bins. The data were aligned with respect to sleep onset, averaged per condition, and plotted against the mean timing of all NREM and REM sleep episodes. Bottom panel: Effect size of the difference between caffeine and placebo expressed as Cohen’s d. Grey dots: p_uncorrected_ < 0.05; black dots: p_uncorrected_ > 0.05. (**B**) Time course of heart rate across the first two NREM-REM sleep episodes in the placebo (green) and caffeine (purple) conditions (mean ± 95% confidence intervals; n = 21). Individual NREM sleep episodes were subdivided into 20 and individual REM sleep episodes into 4 equal time bins. The data were aligned with respect to sleep onset, averaged per condition, and plotted against the mean timing of all NREM and REM sleep episodes. Bottom panel: Effect size of the difference between caffeine and placebo expressed as Cohen’s d. Red dots: p_FDR_ < 0.05; grey dots: p_uncorrected_ < 0.05; black dots: p_uncorrected_ > 0.05.

### Concentration-effect relationship between caffeine and reduction in NREM sleep delta activity

To investigate whether the effect of caffeine on delta activity was concentration dependent, the difference between caffeine and placebo in 0.75-2.5 Hz activity in all time bins during the first two NREM sleep episodes was expressed as a function of the caffeine concentration. Computing the linear correlation coefficient between the mean difference in delta activity and the mean caffeine concentration in the first two NREM sleep episodes revealed a negative association (r_Pearson_ = -0.62, p < 0.005). The generalized-additive, non-linear model function revealed a significant distance from the no-effects line for values above 7.35 µmol/l, suggesting a significant impact of caffeine on EEG delta activity in NREM sleep above this value (Fig. 3).

**Figure 3.**
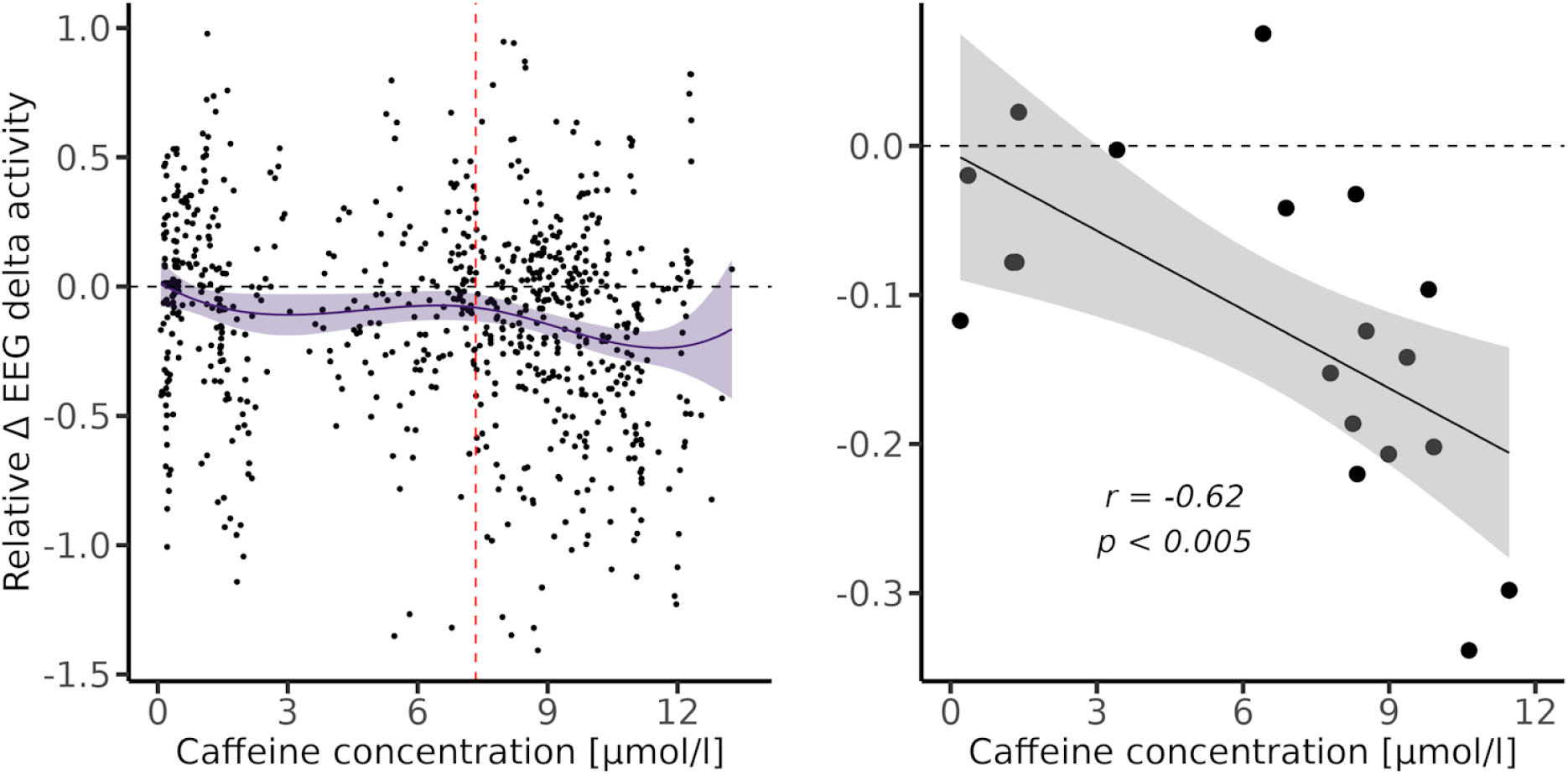
Left panel: Difference in EEG delta activity (0.75-2.5 Hz) between the caffeine and placebo (horizontal dashed line) conditions in 2 x 20 equal time bins of the NREM sleep episodes 1 and 2. Individual data points in 19 participants and their averages (black line ± 95% confidence intervals) are plotted. The red vertical dashed line at 7.34 µmol/l marks the threshold above which the average difference between caffeine and placebo was estimated to be higher than three times the standard error of the null hypothesis. Right panel: Pearson product moment correlation between the difference in EEG delta activity (0.75-2.5 Hz) between caffeine and placebo and the mean caffeine in plasma in the first two NREM sleep episodes (n = 19). The continuous black line shows the corresponding linear trend.

### Effects of caffeine on heart rate during sleep

The time course of heart rate in the placebo and caffeine conditions across the first two NREM-REM sleep cycles is illustrated in Fig. 2B. After a small drop immediately upon sleep onset, heart rate remained low in NREM sleep and transiently increased in REM sleep (*‘state’*, NREM *vs.* REM sleep: F_1,60_ = 58.79, p < 0.0001). Heart rate slightly increased from the 1^st^ to the 2^nd^ NREM sleep episode (F_1,60_ = 4.11, p < 0.05), while it did not differ between the episodes in REM sleep (F_1,34.3_ = 0.17, p = 0.68).

The modulation of heart rate by the different states of vigilance and caffeine is illustrated in Fig. 4. Mean heart rate differed among wakefulness and sleep (*‘state’*: F_4,180_ = 47.12, p < 0.0001), with the lowest values in NREM sleep. Compared to placebo, caffeine induced an overall reduction in heart rate by 3.24 ± 0.77 (SEM) beats-per-minute (BPM) (F_1,180_ = 64.79, p < 0.0001). This difference corresponded to a large effect size (Cohen’s d = 0.79 ± 0.02). In NREM sleep, caffeine reduced heart rate by 3.19 ± 0.78 BPM (F_1,60_ = 36.51, p < 0.0001; N1: t_20_ = 4.03; N2: t_20_ = 4.42; N3: t_20_ = 4.21; p_all_ = 0.001), which corresponded to a large effect size (Cohen’s d = 0.79 ± 0.03). The reduction in REM sleep equaled 3.72 ± 1.01 BPM (F_1,34.3_=22.16, p<0.0001; t_20_ = 2.73, p = 0.012), also corresponding a large effect size (Cohen’s d = 0.8 ± 0.07). In wakefulness, the difference between the conditions was not significant (t_20_ = 1.67, p > 0.1).

**Figure 4.**
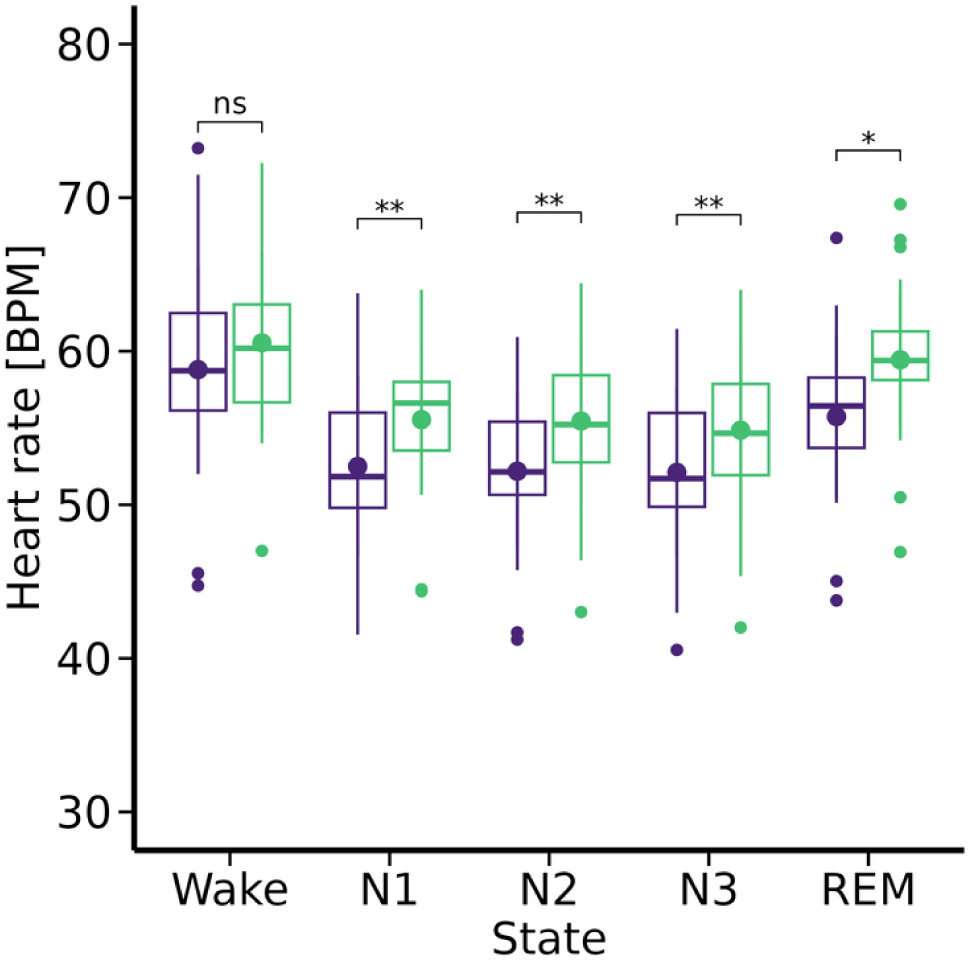
Heart rate in wakefulness (wake), NREM and REM sleep states in the caffeine (purple) and placebo (green) conditions. The box plots represent beats-per-minute (BPM) in 21 study participants. Box: 25^th^, 50^th^ (median) and 75^th^ percentiles; whiskers: 1.5 times the inter-quantile range; dots: individual data points outside the whisker range. Asterisks: two-sided paired t-tests between the conditions: ** p < 0.005, * p < 0.05.

### Concentration-effect relationship between caffeine and reduction in heart rate in NREM sleep

Correlation analysis of the averaged values per participant (i.e., the average of the 2 x 20 time-bin values in NREM sleep) showed a negative correlation between the change of heart rate and the caffeine concentration (r_Pearson_ = -0.66, p < 0.003). Generalized additive model estimates revealed a significant distance from the null-effects line (no difference) above caffeine amounts of 4.25 µmol/l, suggesting that a caffeine concentration above this value reduces heart rate (Fig. 5A).

**Figure 5.**
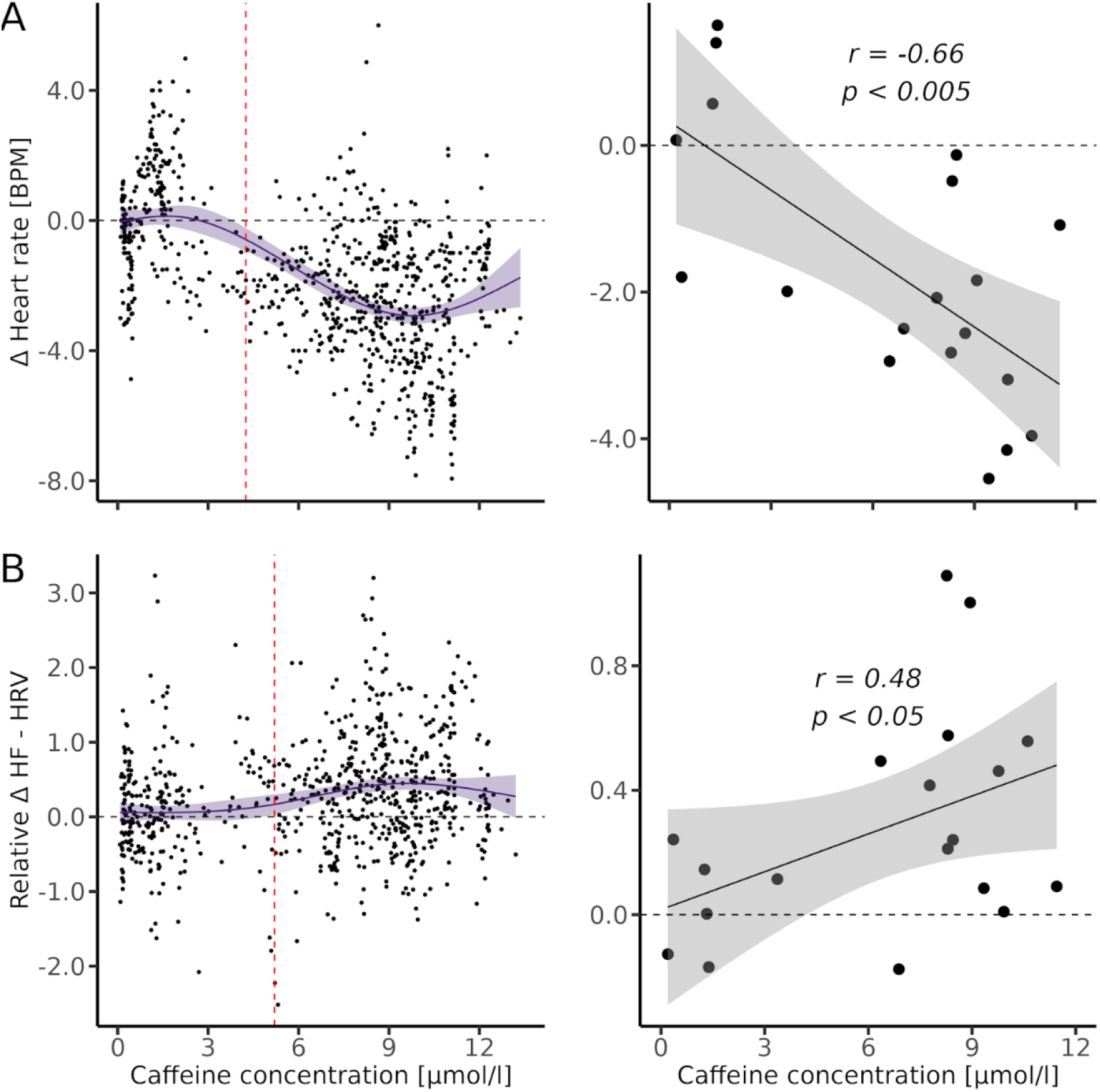
(**A**) Left panel: Difference in heart rate (beats-per-minute, BPM) between the caffeine and placebo (horizontal dashed line) conditions in 2 x 20 equal time bins of the NREM sleep episodes 1 and 2. Individual data points in 19 participants and their averages (black line ± 95% confidence intervals) are plotted. The red vertical dashed line at 4.25 µmol/l marks the threshold above which the average difference between caffeine and placebo was estimated to be higher than three times the standard error of the null hypothesis. Right panel: Pearson product moment correlation between the difference in heart rate between caffeine and placebo and the mean caffeine in plasma in the first two NREM sleep episodes (n = 19). The continuous black line shows the corresponding linear trend. (**B**) Same representation of high-frequency (0.15-0.4 Hz) spectral power in heart rate variability (HR-HRV) as in panel A. The red vertical dashed line at 4.92 µmol/l marks the threshold above which the average difference between caffeine and placebo was estimated to be higher than three times the standard error of the null hypothesis.

### Concentration-effect relationship between caffeine and heart rate variability in NREM sleep

To estimate caffeine-related changes of parasympathetic activity during sleep, we quantified HF-HRV. Caffeine increased the HF-HRV component in NREM sleep by 0.23 ± 0.08 log ms^2^ (F_1,20_ = 7.85, p < 0.05, t_20_ = 2.80, p < 0.05, Cohen’s d = 0.25). Mean individual changes in HF-HRV in the first two NREM sleep episodes correlated positively with the increasing plasma caffeine concentration (r_Pearson_ = 0.48, p < 0.05). The non-linear analysis of the caffeine related change in HF-HRV showed a deviation from the null-effects baseline for caffeine concentrations exceeding 4.92 µmol/l (Fig. 5B).

## Discussion

In this study, we systematically investigated the concentration-effect relationships among the caffeine levels in plasma, the sleep EEG, heart rate, and HF-HRV during human sleep. Using powerful, generalized additive models to probe non-linear associations among these variables, we found concentration-dependent alterations in EEG 0.75-2.5 Hz activity above ∼ 7.3 µmol/l and in heart rate and HF-HRV above ∼ 4-5 µmol/l plasma caffeine. The effects were generally characterized by a consistent and large effect size. The findings suggest that above a certain threshold, caffeine reduces EEG delta activity and increases cardiac autonomic activity in NREM sleep in a concentration-dependent manner. The exact modes of action how caffeine affects these central and autonomic nervous system functions are complex and remain to be fully elucidated.

Sleep architecture, the sleep EEG and cardiac autonomic activity are highly sensitive to subtle internal and external influences such as moderate doses of caffeine. When previous studies aimed at elucidating the concentration- and exposure-effect relationships between caffeine pharmacokinetics and sleep physiologic measures, they typically relied on the caffeine content in a single saliva or blood sample before sleep initiation. Nevertheless, uptake, metabolism and excretion of exogenous caffeine are dynamic processes, which differ widely among individuals. The simultaneous quantification of pharmacokinetic and physiological variables is necessary to understand how they are mutually related. By doing so and capitalizing from the comprehensive characterization of a newly developed, time-controlled, pulsatile-release caffeine formula (Dornbierer et al. 2021), we observed that a caffeine plasma level above 7.3 µmol/l is required to reduce EEG 0.75-2.5 Hz activity in NREM sleep. Above this concentration, we found a robust negative association between this physiological marker of NREM sleep intensity and the level of caffeine.

In humans, the actions of caffeine on the central nervous system are more often quantified by behavioral changes than by direct changes of brain electrical activity. Early experimental work concluded that intake of 80-100 mg caffeine shortly before bedtime is required, to delay the onset of sleep (reviewed by Dews 1982). Furthermore, based on pharmacokinetic estimates, it was suggested that a plasma concentration of ∼ 7 µmol/l (> 2.5 µg/ml) is the threshold for the effective promotion of vigilance during prolonged wakefulness (Beaumont et al. 2001). Interestingly, the present data indicate that a plasma caffeine concentration in this range (∼ 7.3 µmol/l) is necessary to attenuate EEG delta activity in NREM sleep. Given that the saliva/plasma concentration ratio is stable at ∼ 0.74 (Newton et al. 1981), 7 µmol/l caffeine in plasma would correspond to ∼ 10 µmol/l caffeine in saliva. Caffeine intake in the morning and during prolonged wakefulness was repeatedly found to attenuate subsequent EEG slow-wave activity during sleep at concentrations far below this threshold (Landolt et al. 1995; Landolt et al. 2004; Rétey et al. 2007). As discussed in more detail elsewhere, these caffeine-induced changes could reflect the attenuated build-up of homeostatic sleep pressure during wakefulness or the continued antagonism of adenosine receptors by active caffeine metabolites such as paraxanthine (Reichert et al. 2022).

Consistent with previous studies in wakefulness, we found that moderate caffeine decreased heart rate and increased HRV, and that caffeine affects these measures of cardiac autonomic activity in non-linear fashion (Crooks et al. 2019; Kohler at al. 2006; for systematic review, see Koenig et al. 2013). Non-linear statistical analyses revealed that above a plasma concentration of ∼ 4.3 µmol/l, caffeine reduced heart rate on average by more than 3 bpm and that the decrease correlated with increased caffeine levels. Compared to placebo, the heart rate was reduced to a similar extent in all sleep states and wakefulness (we found no significant *‘state’* x *‘condition’* interaction). It was previously hypothesized that low and high doses of caffeine independently affect cardiovascular variables via different underlying mechanisms (Fredholm et al. 2017). While the effects of low-dose caffeine such as in this study may be mediated by adenosine receptor blockade, caffeine at higher doses may induce increased intracellular calcium, norepinephrine release or sensitization of dopamine receptors (Fredholm et al. 2017; Temple et al. 2017). The present research does not allow to elucidate the physiological mechanism underpinning the reduced heart rate. We speculate that it could reflect a baroreflex-induced bradycardia caused by caffeine-induced rise in blood pressure (de Zambotti et al. 2018). Baroreflexes regulate blood pressure, heart rate, and blood volume within a narrow range by activating baroreceptors located in major arteries, veins, and the heart. These stretch-activated receptors signal to the nucleus of the solitary tract of the brain stem through the vagus and glossopharyngeal nerves. They evoke reflex inhibition of sympathetic signals to blood vessels, causing vasodilatation, and increase parasympathetic nerve activity to the sinoatrial node, slowing heart rate (Kaufmann et al., 2020). The exact response of the baroreflex and the autonomic nervous system can vary depending on the sleep stage, stress level and overall health status (Penzel et al. 2016).

To estimate vagally-mediated, parasympathetic nervous system activity, we quantified the HF-HRV during sleep (Koenig et al. 2013; de Zambotti et al. 2018). Compared to placebo, we found that this measure was increased for caffeine levels above ∼4.9 µmol/l, and the increase correlated with increased caffeine levels. Assuming normal sinus rhythm and atrioventricular functions, increased HF-HRV may reflect the parasympathetic modulation of normal R-R intervals driven by ventilation (Kleiger, Stein, and Bigger Jr. 2005). Because we did not quantify the evolution of blood pressure and breathing patterns during sleep, future studies are necessary to investigate this hypothesis. In addition, we don’t know whether the caffeine metabolites paraxanthine, theophylline and theobromine contributed to the observed effect. The metabolic breakdown of caffeine underlies high inter-individual variation and each of the major metabolites affect adenosine receptors in body organs with different affinity and via different downstream mechanisms. These unknown secondary effects of caffeine may contribute to the margin of error, which is evident in the heterogeneous distribution of some data reported.

## Author contributions

DD and HPL conceived and designed the study; DD acquired the data; DMB analyzed the data, prepared the figures and drafted the first version of the manuscript; DMB, DD and HPL revised, edited and approved the manuscript.

## Data Availability

The data produced in the present study are available upon reasonable request to the authors.

## Acknowledgements

We thank Rafael Wespi, Laura Schnider and Gina Elrod for their help with the data acquisition.

## References

1. Beaumont, M., D. Batejat, C. Pierard, O. Coste, P. Doireau, P. Van Beers, F. Chauffard, D. Chassard, M. Enslen, J. B. Denis, and D. Lagarde. 2001. “Slow Release Caffeine and Prolonged (64-h) Continuous Wakefulness: Effects on Vigilance and Cognitive Performance.” Journal of Sleep Research 10(4):265–76. doi: 10.1046/j.1365-2869.2001.00266.x.

2. Berntson, Gary G., J. Thomas Bigger Jr., Dwain L. Eckberg, Paul Grossman, Peter G. Kaufmann, Marek Malik, Haikady N. Nagaraja, Stephen W. Porges, J. Philip Saul, Peter H. Stone, and Maurots W. Van Der Molen. 1997. “Heart Rate Variability: Origins, Methods, and Interpretive Caveats.” Psychophysiology 34(6):623–48. doi: 10.1111/j.1469-8986.1997.tb02140.x.

3. Berry, Richard B., Rita Brooks, Charlene Gamaldo, Susan M. Harding, Robin M. Lloyd, Stuart F. Quan, Matthew T. Troester, and Bradley V. Vaughn. 2017. “AASM Scoring Manual Updates for 2017 (Version 2.4).” Journal of Clinical Sleep Medicine 13(5):665–66. doi: 10.5664/jcsm.6576.

4. Blume, Christine, and Christian Cajochen. 2021. “‘SleepCycles’ Package for R - A Free Software Tool for the Detection of Sleep Cycles from Sleep Staging.” MethodsX 8:101318. doi: 10.1016/j.mex.2021.101318.

5. Bonati, Maurizio, Roberto Latini, Ferruccio Galletti, John F. Young, Gianni Tognoni, and Silvio Garattini. 1982. “Caffeine Disposition after Oral Doses.” Clinical Pharmacology & Therapeutics 32(1):98–106. doi: 10.1038/clpt.1982.132.

6. Carrier, Julie, Jean Paquet, Marta Fernandez-Bolanos, Laurence Girouard, Joanie Roy, Brahim Selmaoui, and Daniel Filipini. 2009. “Effects of Caffeine on Daytime Recovery Sleep: A Double Challenge to the Sleep–Wake Cycle in Aging.” Sleep Medicine 10(9):1016–24. doi: 10.1016/j.sleep.2009.01.001.

7. Clark, Ian, and Hans Peter Landolt. 2017. “Coffee, Caffeine, and Sleep: A Systematic Review of Epidemiological Studies and Randomized Controlled Trials.” Sleep Medicine Reviews 31:70–78. doi: 10.1016/j.smrv.2016.01.006.

8. Cohen, Jacob. 1988. Statistical Power Analysis for the Behavioral Sciences. Hillsdale, N.J.: L. Erlbaum Associates.

9. Crooks, Elena, Devon A. Hansen, Brieann C. Satterfield, Matthew E. Layton, and Hans P. A. Van Dongen. 2019. “Cardiac Autonomic Activity during Sleep Deprivation with and without Caffeine Administration.” Physiology & Behavior 210:112643. doi: 10.1016/j.physbeh.2019.112643.

10. Dews, P. B. 1982. “Caffeine.” Annual Review of Nutrition 2(1):323–41. doi: 10.1146/annurev.nu.02.070182.001543.

11. Dornbierer, Dario A., Firat Yerlikaya, Rafael Wespi, Martina I. Boxler, Clarissa D. Voegel, Laura Schnider, Aslihan Arslan, Diego M. Baur, Markus R. Baumgartner, Tina Maria Binz, Thomas Kraemer, and Hans-Peter Landolt. 2021. “A Novel Bedtime Pulsatile-Release Caffeine Formula Ameliorates Sleep Inertia Symptoms Immediately upon Awakening.” Scientific Reports 11(1):19734. doi: 10.1038/s41598-021-98376-z.

12. Drake, Christopher, Timothy Roehrs, John Shambroom, and Thomas Roth. 2013. “Caffeine Effects on Sleep Taken 0, 3, or 6 Hours before Going to Bed.” Journal of Clinical Sleep Medicine 09(11):1195–1200. doi: 10.5664/jcsm.3170.

13. Efron, Bradley, and R. J. Tibshirani. 1993. An Introduction to the Bootstrap. 1st ed. New York: Chapman and Hall/CRC.

14. Feinberg, I., and T. C. Floyd. 1979. “Systematic Trends Across the Night in Human Sleep Cycles.” Psychophysiology 16(3):283–91. doi: 10.1111/j.1469-8986.1979.tb02991.x.

15. Fredholm, Bertil B., Jiangning Yang, and Yingqing Wang. 2017. “Low, but Not High, Dose Caffeine Is a Readily Available Probe for Adenosine Actions.” Molecular Aspects of Medicine 55:20–25. doi: 10.1016/j.mam.2016.11.011.

16. Gardiner, Carissa, Jonathon Weakley, Louise M. Burke, Gregory D. Roach, Charli Sargent, Nirav Maniar, Andrew Townshend, and Shona L. Halson. 2023. “The Effect of Caffeine on Subsequent Sleep: A Systematic Review and Meta-Analysis.” Sleep Medicine Reviews 101764. doi: 10.1016/j.smrv.2023.101764.

17. Green, Peter J., Robert Kirby, and Jerry Suls. 1996. “The Effects of Caffeine on Blood Pressure and Heart Rate: A Review1.” Annals of Behavioral Medicine 18(3):201–16. doi: 10.1007/BF02883398.

18. Kaufmann, Horacio, Lucy Norcliffe-Kaufmann, and Jose-Alberto Palma. 2020. “Baroreflex Dysfunction.” New England Journal of Medicine 382(2):163–78. doi: 10.1056/NEJMra1509723.

19. Kleiger, Robert E., Phyllis K. Stein, and J. Thomas Bigger Jr. 2005. “Heart Rate Variability: Measurement and Clinical Utility.” Annals of Noninvasive Electrocardiology 10(1):88–101. doi: 10.1111/j.1542-474X.2005.10101.x.

20. Koenig, Julian, Marc N. Jarczok, Wolfgang Kuhn, Katharina Morsch, Alexander Schäfer, Thomas K. Hillecke, and Julian F. Thayer. 2013. “Impact of Caffeine on Heart Rate Variability: A Systematic Review.” Journal of Caffeine Research 3(1):22–37. doi: 10.1089/jcr.2013.0009.

21. Kohler, Mark, Alan Pavy, and Cameron Van Den Heuvel. 2006. “The Effects of Chewing versus Caffeine on Alertness, Cognitive Performance and Cardiac Autonomic Activity during Sleep Deprivation.” Journal of Sleep Research 15(4):358–68. doi: 10.1111/j.1365-2869.2006.00547.x.

22. Landolt, Hans-Peter, Derk-Jan Dijk, Stephanie E. Gaus, and Alexander A. Borbély. 1995. “Caffeine Reduces Low-Frequency Delta Activity in the Human Sleep EEG.” Neuropsychopharmacology 12(3):229–38. doi: 10.1016/0893-133X(94)00079-F.

23. Landolt, Hans-Peter, Julia V. Rétey, Karin Tönz, Julie M. Gottselig, Ramin Khatami, Isabelle Buckelmüller, and Peter Achermann. 2004. “Caffeine Attenuates Waking and Sleep Electroencephalographic Markers of Sleep Homeostasis in Humans.” Neuropsychopharmacology 29(10):1933–39. doi: 10.1038/sj.npp.1300526.

24. Landolt, Hans-Peter, Esther Werth, Alexander A. Borbély, and Derk-Jan Dijk. 1995. “Caffeine Intake (200 Mg) in the Morning Affects Human Sleep and EEG Power Spectra at Night.” Brain Research 675(1):67–74. doi: 10.1016/0006-8993(95)00040-W.

25. Lazarus, Michael, Yo Oishi, Theresa E. Bjorness, and Robert W. Greene. 2019. “Gating and the Need for Sleep: Dissociable Effects of Adenosine A1 and A2A Receptors.” Frontiers in Neuroscience 13:740. doi: 10.3389/fnins.2019.00740.

26. Newton, R., L. J. Broughton, M. J. Lind, P. J. Morrison, H. J. Rogers, and I. D. Bradbrook. 1981. “Plasma and Salivary Pharmacokinetics of Caffeine in Man.” European Journal of Clinical Pharmacology 21(1):45–52. doi: 10.1007/BF00609587.

27. Penzel, Thomas, Jan W. Kantelhardt, Ronny P. Bartsch, Maik Riedl, Jan F. Kraemer, Niels Wessel, Carmen Garcia, Martin Glos, Ingo Fietze, and Christoph Schöbel. 2016. “Modulations of Heart Rate, ECG, and Cardio-Respiratory Coupling Observed in Polysomnography.” Frontiers in Physiology 7. doi: 10.3389/fphys.2016.00460.

28. Reichert, Carolin Franziska, Tom Deboer, and Hans-Peter Landolt. 2022. “Adenosine, Caffeine, and Sleep–Wake Regulation: State of the Science and Perspectives.” Journal of Sleep Research 31(4):e13597. doi: 10.1111/jsr.13597.

29. Rétey, J. V., M. Adam, R. Khatami, U. F. O. Luhmann, H. H. Jung, W. Berger, and H. P. Landolt. 2007. “A Genetic Variation in the Adenosine A2A Receptor Gene (ADORA2A) Contributes to Individual Sensitivity to Caffeine Effects on Sleep.” Clinical Pharmacology & Therapeutics 81(5):692–98. doi: 10.1038/sj.clpt.6100102.

30. Riksen, Niels P., Paul Smits, and Gerard A. Rongen. 2011. “The Cardiovascular Effects of Methylxanthines.” Pp. 413–37 in Methylxanthines, Handbook of Experimental Pharmacology, edited by B. B. Fredholm. Berlin, Heidelberg: Springer.

31. Temple, Jennifer L., Christophe Bernard, Steven E. Lipshultz, Jason D. Czachor, Joslyn A. Westphal, and Miriam A. Mestre. 2017. “The Safety of Ingested Caffeine: A Comprehensive Review.” Frontiers in Psychiatry 8:80. doi: 10.3389/fpsyt.2017.00080.

32. Turnbull, Duncan, Joseph V. Rodricks, Gregory F. Mariano, and Farah Chowdhury. 2017. “Caffeine and Cardiovascular Health.” Regulatory Toxicology and Pharmacology 89:165–85. doi: 10.1016/j.yrtph.2017.07.025.

33. Van Dongen, Hans P. A., Nicholas J. Price, Janet M. Mullington, Martin P. Szuba, Shiv C. Kapoor, and David F. Dinges. 2001. “Caffeine Eliminates Psychomotor Vigilance Deficits from Sleep Inertia.” Sleep 24(7):813–19. doi: 10.1093/sleep/24.7.813.

34. Whitsett, Thomas L., Carl V. Manion, and H. Dix Christensen. 1984. “Cardiovascular Effects of Coffee and Caffeine.” The American Journal of Cardiology 53(7):918–22. doi: 10.1016/0002-9149(84)90525-3.

35. de Zambotti, Massimiliano, John Trinder, Alessandro Silvani, Ian Colrain, and Fiona C. Baker. 2018. “Dynamic Coupling between the Central and Autonomic Nervous Systems during Sleep: A Review.” Neuroscience and Biobehavioral Reviews 90:84–103. doi: 10.1016/j.neubiorev.2018.03.027.

