## Supplemental material. for "Concentration-effect relationships of plasma caffeine on EEG delta power and cardiac autonomic activity during human sleep"

**Running head:** Concentration-effect relationships of caffeine on the EEG and heart rate during sleep

**Submitted to:** *The Journal of Sleep Research*
12 October 2023

### Supplemental information

**Address for correspondence:**
Prof. Hans-Peter Landolt, PhD
Institute of Pharmacology & Toxicology
University of Zürich
Winterthurerstrasse 190
8057 Zürich, Switzerland


**Supplementary Table S1.** Visually scored sleep variables during the first two NREM-REM sleep cycles.

| | Placebo | | Caffeine | | | | |
|---|---|---|---|---|---|---|---|
| **Variable** | **Mean** | **SEM** | **Mean** | **SEM** | **t-value** | **df** | **$p_{FDR}$** |
| TST | 153.78 | 5.79 | 142.84 | 6.71 | -1.70 | 20 | 0.2297 |
| SL | 11.93 | 4.38 | 13.81 | 3.63 | 0.82 | 20 | 0.5500 |
| RL | 70.26 | 11.08 | 87.67 | 11.58 | 1.11 | 20 | 0.4030 |
| N1 | 4.95 | 0.84 | 7.62 | 0.92 | 2.57 | 20 | 0.1014 |
| N2 | 56.19 | 3.84 | 55.07 | 2.71 | -0.30 | 20 | 0.7660 |
| N3 | 69.67 | 4.27 | 55.76 | 4.26 | -3.57 | 20 | **0.0250** |
| REM sleep | 18.81 | 4.00 | 20.64 | 3.44 | 0.42 | 20 | 0.7334 |
| WASO | 5.78 | 2.00 | 10.9 | 3.83 | 1.27 | 20 | 0.3575 |

Mean values are reported in minutes (± the standard error of the mean [SEM]) for 21 study participants. Total sleep time (TST): time spent in sleep stages N1, N2, N3 and REM sleep between sleep onset and the end of the second REM sleep episode. Sleep latency (SL): time between lights-off and first occurrence of stage N2. REM sleep latency (RL): time between lights-off and first occurrence of REM sleep. N1, N2, N3: non-rapid-eye-movement sleep stages. REM sleep: rapid-eye-movement sleep. WASO: wakefulness after sleep onset. Paired t-test was used for the comparisons between the conditions; false discovery rate (FDR) was used to correct for multiple comparison.

**Supplementary Figure S1.**

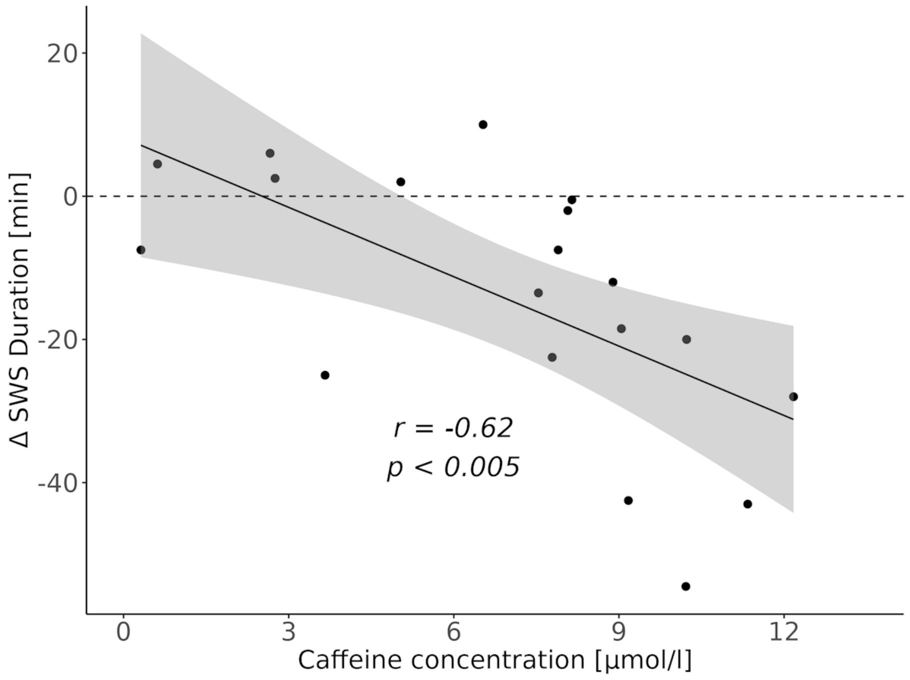


Pearson product moment correlation between the difference in slow wave sleep (min) between the caffeine and placebo conditions, and the mean caffeine concentration in blood plasma in the first two NREM sleep episodes. The continuous black line illustrates the corresponding linear trend ($r = -0.62$, $p < 0.005$, $n = 19$).
